# The efficacy of sampling strategies for estimating scabies prevalence

**DOI:** 10.1101/2021.11.13.21266293

**Authors:** Nefel Tellioglu, Rebecca H. Chisholm, Jodie McVernon, Nicholas Geard, Patricia T. Campbell

## Abstract

**Background:** Estimating scabies prevalence in communities is crucial for identifying the communities with high scabies prevalence and guiding interventions. There is no standardisation of sampling strategies to estimate scabies prevalence in communities, and a wide range of sampling sizes and methods have been used. The World Health Organization recommends household sampling or, as an alternative, school sampling to estimate community-level prevalence. Due to varying prevalence across populations, there is a need to understand how sampling strategies for estimating scabies prevalence interact with scabies epidemiology to affect accuracy of prevalence estimates.

**Methods:** We used a simulation-based approach to compare the efficacy of different sampling methods and sizes. First, we generate synthetic populations with Australian Indigenous communities’ characteristics and then, assign a scabies status to individuals to achieve a specified prevalence using different assumptions about scabies epidemiology. Second, we calculate an observed prevalence for different sampling methods and sizes.

**Results:** The distribution of prevalence in population groups can vary substantially when the underlying scabies assignment method changes. Across all of the scabies assignment methods combined, the simple random sampling method produces the narrowest 95% confidence interval for all sampling percentages. The household sampling method introduces higher variance compared to simple random sampling when the assignment of scabies includes a household-specific component. The school sampling method overestimates community prevalence when the assignment of scabies includes an age-specific component.

**Discussion:** Our results indicate that there are interactions between transmission assumptions and surveillance strategies, emphasizing the need for understanding scabies transmission dynamics. We suggest using the simple random sampling method for estimating scabies prevalence. Our approach can be adapted to various populations and diseases.

**Author summary:** Scabies is a parasitic infestation that is commonly observed in underprivileged populations. A wide range of sampling sizes and methods have been used to estimate scabies prevalence. With differing key drivers of transmission and varying prevalence across populations, it can be challenging to determine an effective sampling strategy. In this study, we propose a simulation approach to compare the efficacy of different sampling methods and sizes. First, we generate synthetic populations and then assign a scabies status to individuals to achieve a specified prevalence using different assumptions about scabies epidemiology. Second, we calculate an observed prevalence for different sampling methods and sizes. Our results indicate that there are interactions between transmission assumptions and surveillance strategies. We suggest using the simple random sampling method for estimating prevalence as it produces the narrowest 95% confidence interval for all sampling sizes. We propose guidelines for determining a sample size to achieve a desired level of precision in 95 out 100 samples, given estimates of the population size and a priori estimates of true prevalence. Our approach can be adapted to various populations, informing an appropriate sampling strategy for estimating scabies prevalence with confidence.

## Introduction

Scabies is a parasitic infestation caused by the mite *Sarcoptes scabiei* [1] and is one of the highest-burden Neglected Tropical Diseases (NTDs) [2]. In 2016, it was estimated that scabies affects 455 million people annually and causes 3.8 million disability-adjusted life years (DALYs) [2]. The prevalence of scabies is highest in underprivileged tropical settings including Indigenous communities of Australia and Pacific Island communities [3–5]. These settings are thought to be affected due to factors such as overcrowding [6], hot weather and humidity [7, 8]. Scabies prevalence can reach up to 35% in remote Indigenous communities [9, 10] and 71% in Pacific Island communities [11]. In these humid low-income settings, the scratching due to scabies can lead to secondary skin infections by Group A *Streptococcus* and their sequelae, which scales up the burden of scabies [8, 12]. Interventions for controlling scabies can reduce the burden of not only scabies but also the secondary skin infections [8]. Such interventions can be costly to implement [4] and it is desirable to focus such efforts on high prevalence settings.

A recent report of the World Health Organization (WHO) Informal Consultation on a Framework for Scabies Control suggests that an MDA is needed when observed scabies prevalence is more than ten percent [28]. For this intervention to be applied appropriately, an accurate estimation of true prevalence in communities is crucial. For prevalence estimation, WHO recommends community-based household sampling methods including people from all ages as the most appropriate strategy. They also suggest that school-based sampling might be an alternative, however, note further research is needed to determine how school scabies prevalence is related to community prevalence [28]. Even though the WHO recommends sampling strategies for estimating scabies prevalence, they underline the need for evaluation of the efficacy of such strategies. The design of studies that are both efficient and unbiased can be challenging due to following key issues.

First, the extent to which standard sample size calculation formulas are applicable to scabies is uncertain, as substantial heterogeneity in prevalence is observed between households and age groups. Prevalence studies have shown that household contacts play a crucial role in scabies transmission [14, 15]. It is estimated that it takes on average 20 minutes of close contact for scabies transmission [15] which highlights household contacts as an important factor in scabies transmission [15–17]. Dagne et al. [14] found that the probability of being infested by scabies was almost five times higher among participants with at least one household member having an itchy lesion than participants without family members with such a lesion, underlying the crucial role of the household in transmission. Other prevalence studies conducted in different settings have found scabies prevalence to be age-dependent, with children experiencing prevalence around two to three times that of adults [11, 13, 18–22].

Second, it is hard to compare effectiveness of different sampling strategies across populations. Prevalence estimation studies have used different sampling strategies due to the varying reasons such estimates were required (for example estimating the prevalence in schoolchildren or applying mass drug administration (MDA)) [13, 14, 23–27]. For example, to estimate the level of treatment uptake in households with clinically diagnosed scabies cases, La et al.[26] screened households based on previous enrolment into a related study and found that 23% of the screened population had scabies before the intervention. To estimate scabies prevalence in a welfare home in Malaysia, Zayyid et al. [27] screened a random selection of 120 out of 160 children and found 31% of children had scabies. Moreover, highly variable scabies prevalence has been observed in survey studies conducted in Australian Indigenous communities (from 5% to 35%) [11], and other Pacific Island communities (from 5% to 71%) [11]. With no standardization in methods for scabies prevalence estimation [4], it is difficult to make valid comparisons across settings.

In order to evaluate sampling strategies for infectious disease prevalence, there are published simulation-based approaches [29–35]. Such simulation-based approaches allow us to introduce disease and population-specific characteristics and conduct *in silico* experiments on the effectiveness of sampling strategies. For example, Giardina et al. [29] used a dynamic simulation to compare efficacy of sampling strategies for monitoring morbidity targets for soil-transmitted helminths in districts consisting of villages. They found that sampling school-aged children from ten instead of five villages would increase the sampling effectiveness by 20%. Schmidt et al. [30] found that clustering among individuals and infection duration were major factors contributing to the effectiveness of sampling strategies to measure the prevalence of recurrent infections.

With the uncertainty around age-dependent prevalence and level of household transmission, it remains to be determined whether common sampling approaches introduce a bias in estimating the true prevalence of scabies. In this study, we evaluated the efficacy of different sampling strategies to estimate scabies prevalence using a simulation-based approach. We demonstrate our approach in the context of remote Australian Indigenous communities. Our approach allows comparison of the performance of sampling strategies in a simulated population which has similar age and household size distributions to Australian Indigenous communities.

## Methods

In order to compare and evaluate different sampling strategies, we simulated a population with a known true prevalence of scabies, and also simulated the sampling strategy used to estimate the prevalence of scabies in that population ^1^. By comparing the estimated prevalence to the true prevalence, we evaluated the efficacy of a given sampling strategy and compare the performance of different strategies. Our approach consists of three stages. First, we generated synthetic populations with characteristics similar to those of remote Australian Indigenous communities. Second, given uncertainty around the relative importance of age- and household-specific factors, we examined five different rules for attributing disease status in the population. We assigned a positive or negative scabies status to all individuals in the population to achieve a specified prevalence. Third, we sampled a percentage of this synthetic population using a pre-defined sampling strategy and sample size and record the sample prevalence. Finally, we compared the specified prevalence (the input prevalence) with the sample prevalence (the output prevalence).

### Generating synthetic populations

We generated populations ranging in size from approximately 500 to 4000 individuals representing the population size of medium to large remote Australian Indigenous communities [36]. Within each population, we assigned individuals into households and age-classes. Age-classes consist of *adult* (16 years and over), *school* (5–15 years), and *pre-school* (0–4 years) at random such that the household size distribution and age distribution of the population reflected Australian Bureau of Statistics (ABS) 2016 census data [37] and survey data of Vino et al. [38].

To generate a population of size *N*, we repeatedly sampled household sizes from the household size distribution of Indigenous communities in ABS data until the population contained approximately *N* people (with a tolerance of 5%). In ABS data, the distribution of households having a size of six or larger is aggregated. After sampling household sizes ranging [1, …, 5, 6+], we used survey data of Vino et al. [38] to disaggregate data for households with a size of six or larger. Household size distribution of all of the simulated populations (blue) and ABS data (red) are represented in Fig 1.

**Fig 1.**
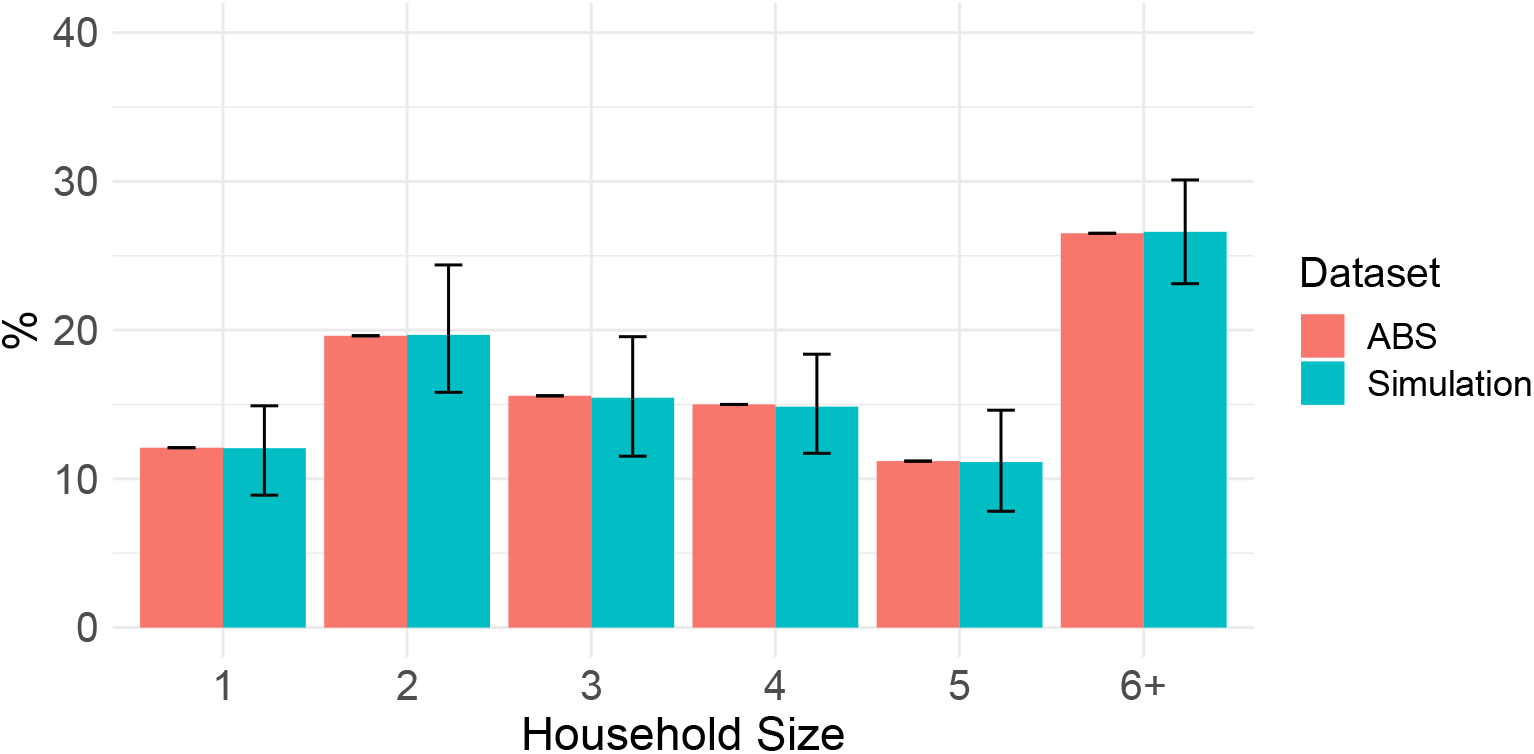
Household size distribution. Household size distribution (median and 2.5-97.5 quantiles) of simulated data (blue) and Indigenous communities household size distribution taken from Australian Bureau of Statistics 2016 census data (red) [37] are presented.

For each household, given a household size, we sampled age-classes (*adult, school, pre-school*) for household members based on age-class distribution of household data from [38]. We repeated the sampling of age-classes for each household until it contained at least one adult. In the survey data, there was no age-class distribution for households of size 15, 18, 19, 20, and 22. For these households, we used the age-class distribution of the closest household size. We then compared the age-class structure of all of the simulated populations with independent age structure data from the Aboriginal and Torres Strait Islander Health Performance Framework 2017 [39] (Fig 2).

**Fig 2.**
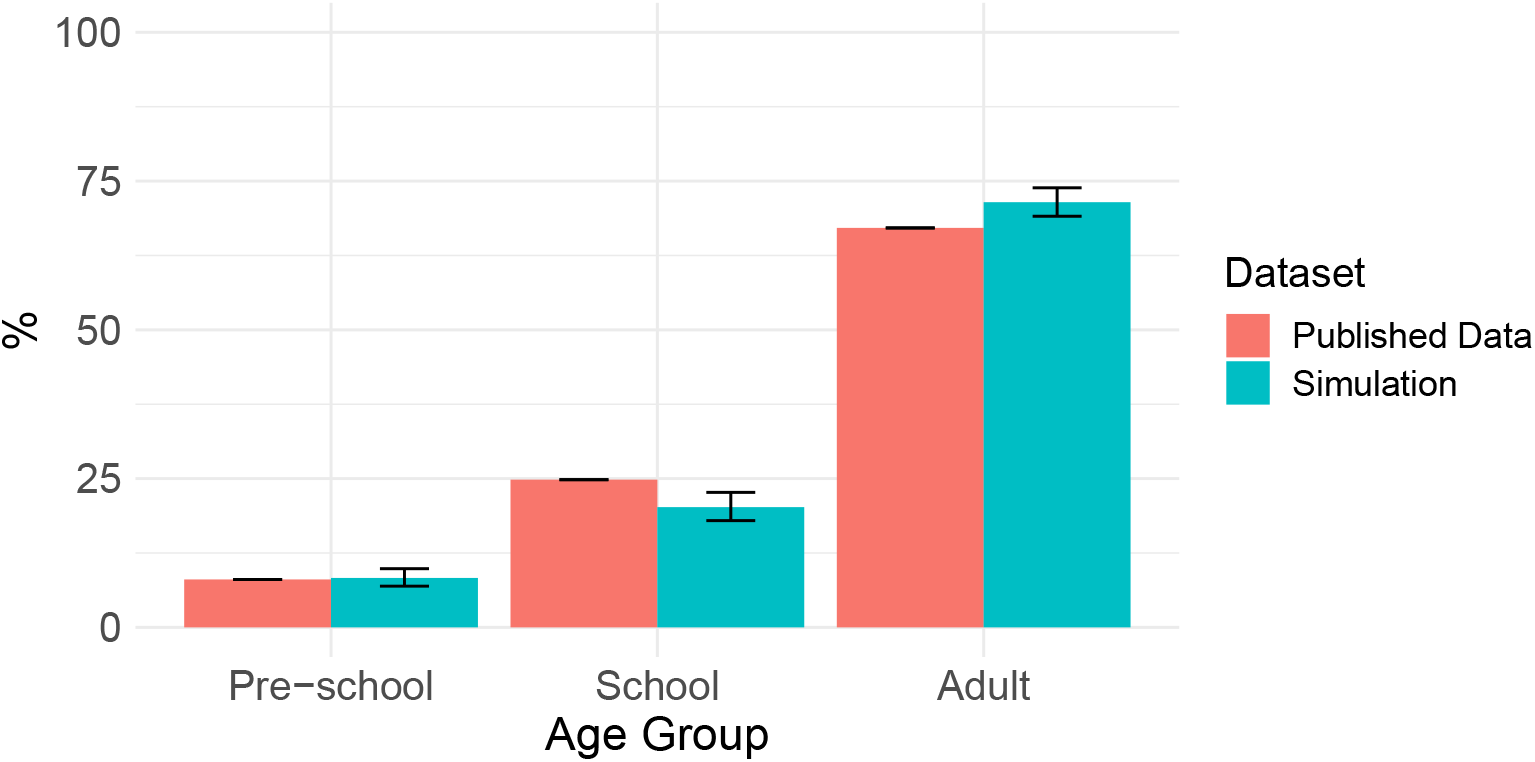
Age distribution. Age distribution (median and 2.5-97.5 quantiles) of simulated data (blue) and age distribution taken from Aboriginal and Torres Strait Islander Health Performance Framework 2017 (red) [39] are presented.

### Assigning scabies status

For each of the generated populations, we assigned a positive or negative scabies status to individuals to achieve an input prevalence percentage, ranging from 5% to 40%. The *assignment method* used to set the scabies status of individuals was chosen from one of the following:

1. **Random:** Individuals were assigned a positive scabies status uniformly at random.
2. **Household-specific (high):** Households were selected uniformly at random, and all individuals in the selected households were assigned a positive scabies status.
3. **Household-specific (mild):** Households were selected uniformly at random, and half of the individuals in the selected households were assigned a positive scabies status, uniformly at random.
4. **Age-specific:** Individuals were assigned a positive scabies status uniformly at random; however, children were three times more likely than adults to be assigned a positive scabies status, based on scabies prevalence surveys conducted in NT, Fiji, and Ethiopia [18–20].
5. **Age-and-household-specific:** Households were selected uniformly at random, and a positive scabies status was assigned to half of the individuals in selected households, with children three times more likely than adults to be assigned a positive scabies status.

### Simulating sampling strategies

We simulated three different *sampling methods* in each of the generated populations: random, household or school sampling with *sample sizes* between 5% and 90% of the population, assuming all individuals were available for sampling. In this study, we refer to a combination of a sampling method and a sampling size as a *sampling strategy*. The simple random sampling method involved sampling individuals uniformly at random. The household sampling method involved selecting households uniformly at random and sampling all members. The school sampling method involved sampling individuals uniformly at random from the school age group only.

### Study design

A population is generated with a size sampled uniformly at random in the range of 500 to 4000. For each generated population with age and household structure, we assign scabies with one of the five assignment methods followed by sampling with the chosen strategy (Table 1). For school sampling, where it is not possible to sample the chosen sample size due to the size of the schoolchildren population, we stop sampling when all schoolchildren have been selected. This process is repeated 500 times. A simplified pseudo code is provided in Fig 3.

**Table 1.**
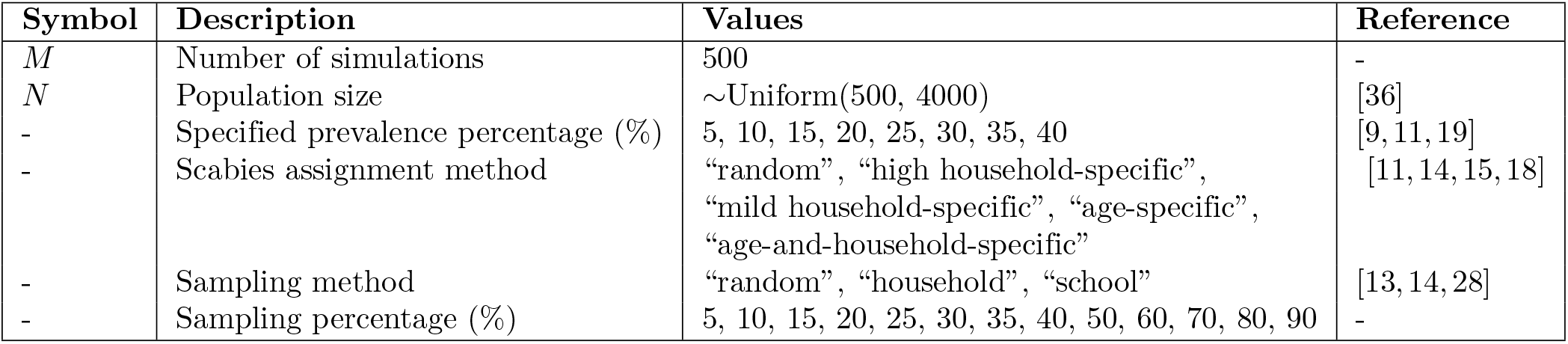
Model Parameters.

| Symbol | Description | Values | Reference |
| --- | --- | --- | --- |
| $M$ | Number of simulations | 500 | - |
| $N$ | Population size | $\sim \text{Uniform}(500, 4000)$ | [36] |
| - | Specified prevalence percentage (%) | 5, 10, 15, 20, 25, 30, 35, 40 | [9, 11, 19] |
| - | Scabies assignment method | “random”, “high household-specific”, “mild household-specific”, “age-specific”, “age-and-household-specific” | [11, 14, 15, 18] |
| - | Sampling method | “random”, “household”, “school” | [13, 14, 28] |
| - | Sampling percentage (%) | 5, 10, 15, 20, 25, 30, 35, 40, 50, 60, 70, 80, 90 | - |

**Fig 3.**
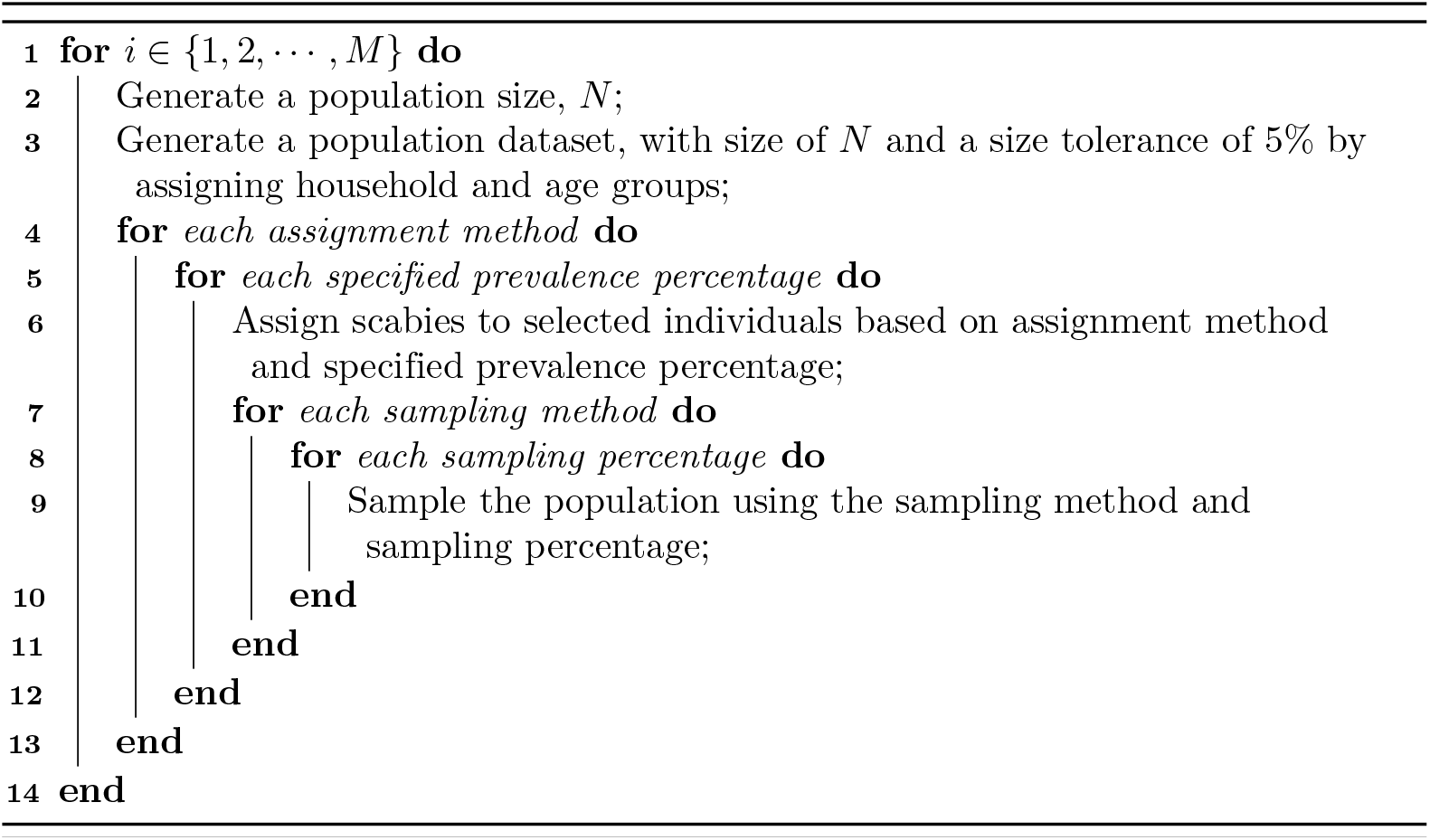
Pseudo code of our algorithm.

In order to compare different sampling methods, we first calculated scabies prevalence distribution in age and household groups given a scabies assignment method and input prevalence. Then, as an exemplar, we compared the output prevalence in the samples using different sampling strategies when the input prevalence was between 20–30%. Finally, we calculated the sample size required to achieve a target precision for each sampling method under different population size, prevalence, and assignment method scenarios.

## Results

We observed that the distribution of prevalence in population groups can vary substantially when the underlying scabies assignment method changes. For example, age-specific scabies assignment increases the prevalence among children as well as prevalence in larger households, due to the higher number of children in larger households (Fig 4 & 5). In addition, household specific assignment approaches introduce higher variance in prevalence among households (Fig 6).

**Fig 4.**
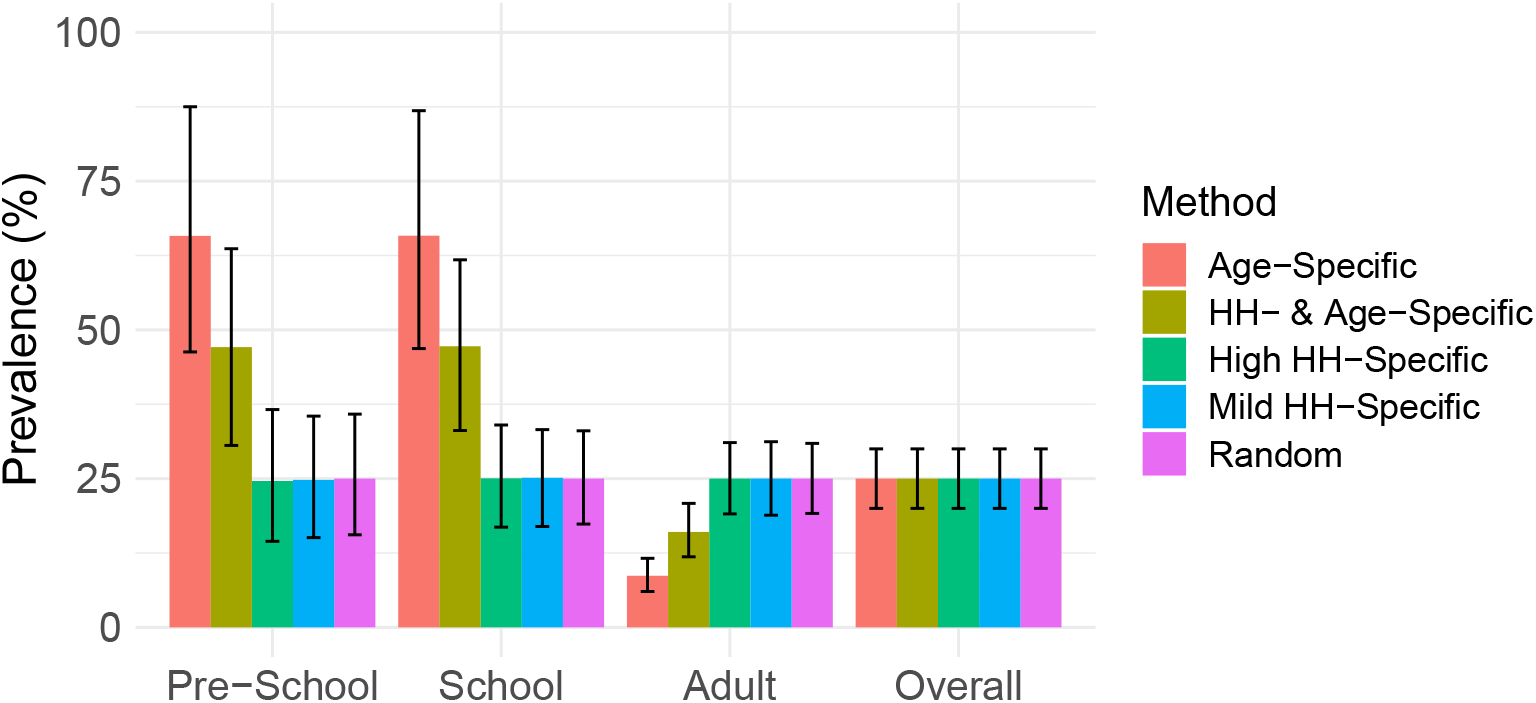
Distribution of scabies prevalence in age groups for different scabies assignment methods. The results (median and 2.5% to 97.5% quantiles) are plotted for an exemplar input prevalence percentage between 20–30% across all population sizes.

**Fig 5.**
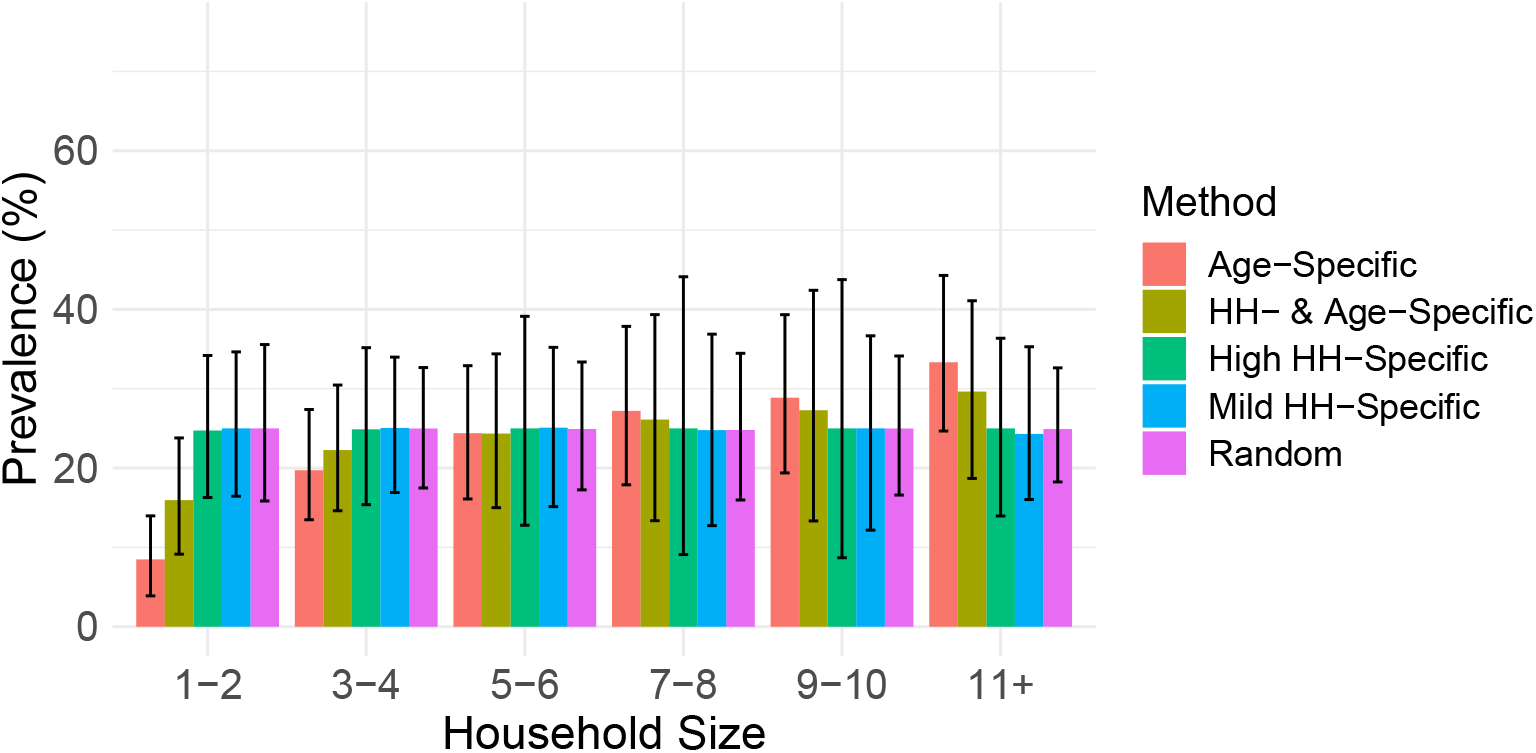
Distribution of scabies prevalence across household size groups for different methods of scabies status assignment. The results (median and 2.5% to 97.5% quantiles) are plotted for an exemplar input prevalence percentage between 20–30% across all population sizes.

**Fig 6.**
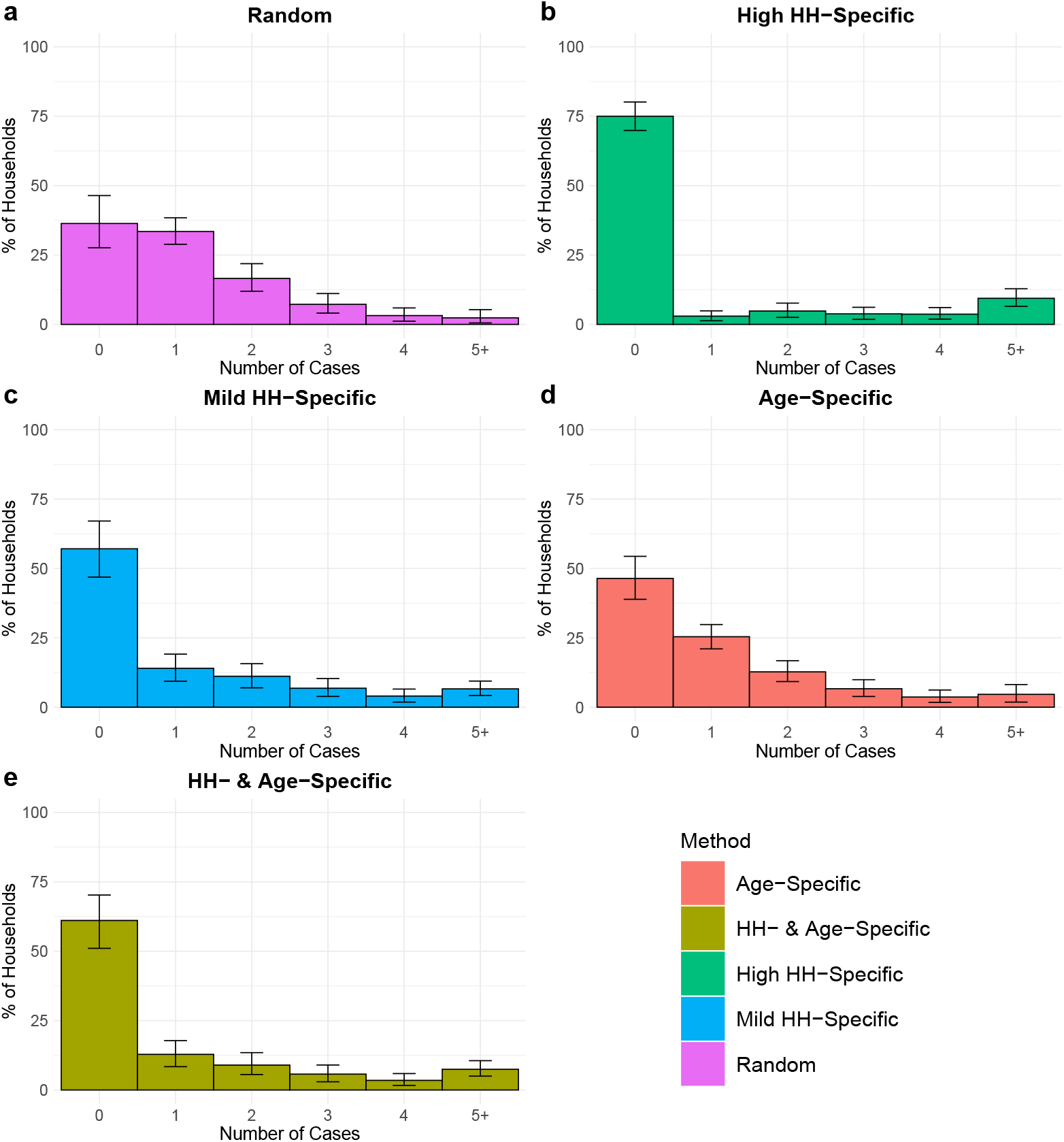
The percentage of households where there are 0, 1, 2, 3, 4, 5+ cases across the different methods of scabies status assignment. The results (median and 2.5% to 97.5% quantiles) are plotted for an exemplar input prevalence percentage between 20–30% across all population sizes.

In Fig 7, we present how the efficiency of sampling methods changes in response to different underlying scabies assignment approaches for an exemplar sampling percentage and input prevalence between 20–30%. The school sampling strategy overestimates the prevalence when the assignment of scabies includes an age-specific component. In addition, the household sampling strategy introduces higher variance compared to simple random sampling when the assignment of scabies includes a household-specific component, because the households with scabies can be over- or under-selected in the samplings. Across all of the scabies assignment methods combined, the simple random sampling strategy produces the narrowest 95% confidence interval for all sampling percentages (Fig 8). The dependence of observed prevalence in the samples on the underlying scabies assignment approach remains across different sampling percentages (S1 Fig).

**Fig 7.**
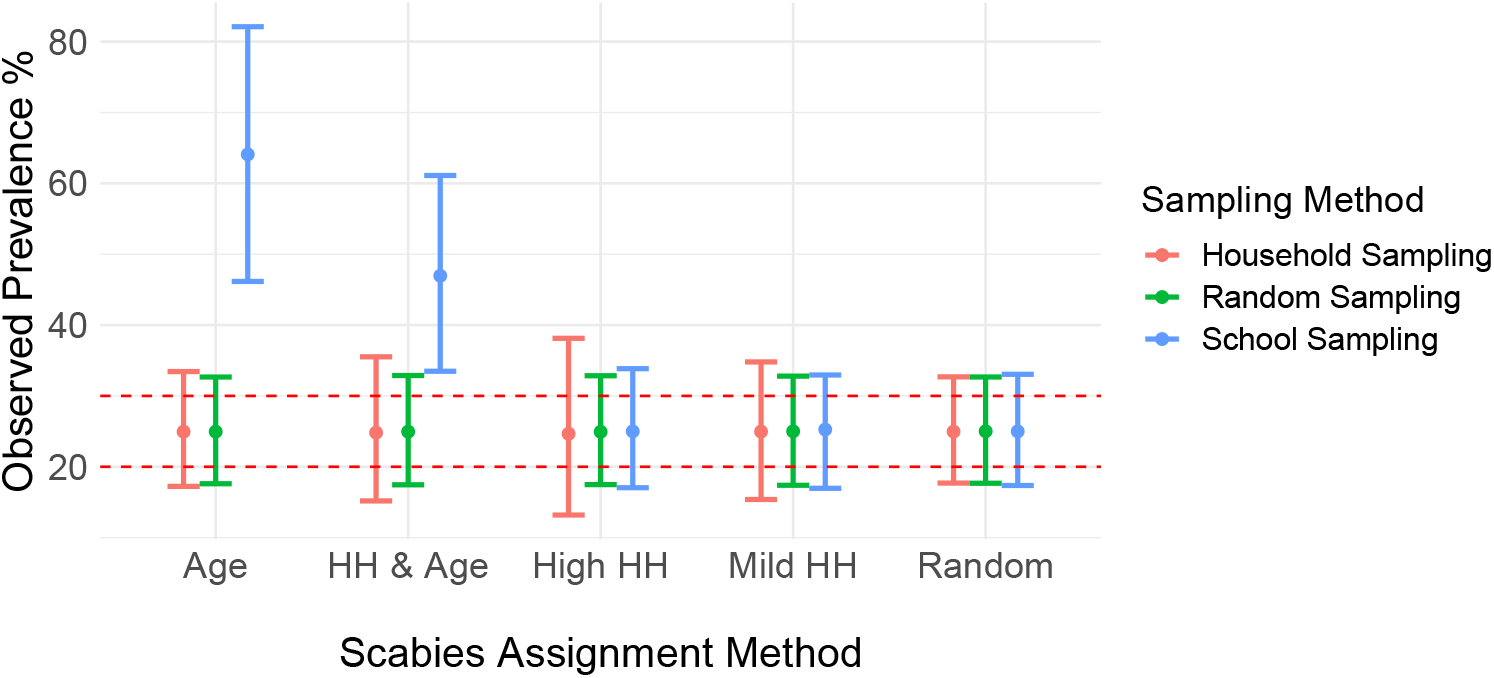
Observed scabies prevalence in samples selected using different sampling methods. The results (median and 2.5% to 97.5% quantiles) are plotted for an exemplar input prevalence percentage between 20–30% across all population sizes with a sampling percentage between 20–30%. Red dashed lines represent 20% and 30% prevalence. Additional results with differing input prevalence and differing population sizes are presented in S2 Fig.

**Fig 8.**
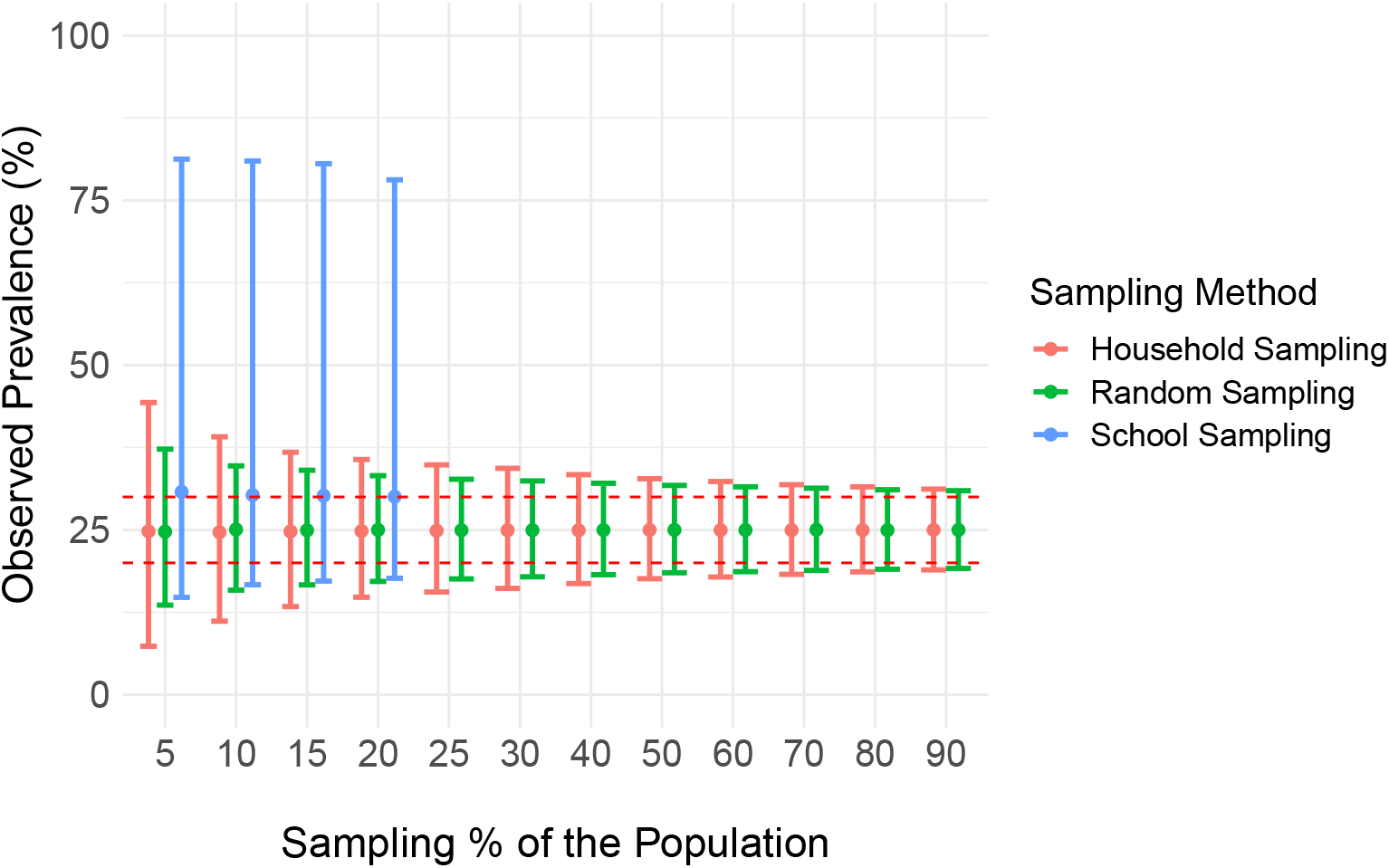
Observed scabies prevalence in samples selected using different sampling methods and sampling percentages. The results (median and 2.5% to 97.5% quantiles) are plotted for an exemplar input prevalence percentage between 20–30% across all population sizes with a sampling percentage between 20–30%. Red dashed lines represent 20% and 30% prevalence. Additional results with differing population sizes and differing input prevalences are presented in S3 Fig and S4 Fig. In the school-based sampling strategy the highest sampling percentages could not be achieved due to insufficient population size in the school aged group.

Table 2 shows the percentage of our synthetic population that needed to be sampled to achieve sample prevalence within the window of input prevalence +/- a stated precision level in 95% of simulations. This table can be used to estimate required sample sizes in real-world populations, based on the population size, an *a priori* estimate of true population prevalence and a desired precision level. For example, for a small population of between 500 and 1500 individuals and an *a priori* estimate of true prevalence between 10–20%, it is necessary to randomly sample 25% of the population to achieve “true prevalence +/- 5%” in 95 out of 100 samples.

**Table 2.** Required sample sizes estimated from simulation results for the simple random, household, and school sampling methods to achieve a given precision, combined across all scabies assignment methods.

| Population Size | *a priori* prevalence (%) | Precision |  |  |  |  |  |  |  |  |
| --- | --- | --- | --- | --- | --- | --- | --- | --- | --- | --- |
|  |  | Simple Random Sampling |  |  | Household Sampling |  |  | School Sampling |  |  |
|  |  | 2% | 5% | 10% | 2% | 5% | 10% | 2% | 5% | 10% |
| Small | 5–10 | 60% | 15% | 5% | 90% | 40% | 15% | X | X | X |
| Small | >10–20 | 70% | 25% | 10% | >90% | 50% | 20% | X | X | X |
| Small | >20–30 | 80% | 30% | 10% | >90% | 60% | 25% | X | X | X |
| Small | >30–40 | >90% | 40% | 15% | >90% | 70% | 30% | X | X | X |
| Medium | 5–10 | 40% | 10% | 3% | 70% | 25% | 10% | X | X | X |
| Medium | >10–20 | 50% | 15% | 3% | 80% | 40% | 10% | X | X | X |
| Medium | >20–30 | 70% | 20% | 5% | 90% | 40% | 15% | X | X | X |
| Medium | >30–40 | >90% | 20% | 5% | >90% | 50% | 15% | X | X | X |
| Large | 5–10 | 25% | 5% | 3% | 60% | 15% | 5% | X | X | X |
| Large | >10–20 | 40% | 10% | 3% | 70% | 25% | 10% | X | X | X |
| Large | >20–30 | 50% | 10% | 3% | 80% | 30% | 10% | X | X | X |
| Large | >30–40 | >90% | 15% | 3% | >90% | 40% | 10% | X | X | X |

Small, medium, and large population sizes represent ranges of [500, 1500], (1500, 2500], (2500, 4000], respectively. For the scenarios with X's in school sampling, sampling all school-aged children was insufficient to have 95% confidence that prevalence within the selected precision could be obtained. Results stratified by scabies assignment methods are shown in S1 Table, S2 Table, S3 Table, S4 Table, S5 Table.

We did not run simulations with a sampling percentage higher than 90%. For the scenarios with “> 90%” in Table 2, sampling 90% of the population was insufficient to have 95% confidence that prevalence within the selected precision could be obtained. Therefore, we do not report the sample size required for these scenarios.

Table 2 shows that the required sampling percentage of the population (1) increases when greater precision is needed, (2) increases with a higher *a priori* prevalence, and (3) decreases with a larger population.

## Discussion

In this study, we present a method to test efficacy of common sampling strategies for scabies in the context of remote Indigenous communities of Australia. To the best of our knowledge, this study is the first to use a simulation-based approach to test the efficacy of scabies sampling strategies, meeting a critical need identified by the WHO [28]. In this section, we discuss how the performance of sampling strategies depends on our assumptions about the relative importance of household- and age-specific scabies transmission, how our analysis can be used in determining sampling size. Then, we provide the strengths and limitations of our study and our future work.

Our results demonstrate how the performance of sampling methods strongly depends on the underlying drivers of scabies transmission, due to the substantial changes in the distribution of scabies prevalence across population groups depending on how scabies spreads. As the precise drivers of scabies distribution within populations are unknown [11, 13–15, 18–20], we cannot be sure which of the scabies assignment methods we have used is closest to reality. Therefore, it is important to use a sampling approach that performs well across all the scabies assignment methods. Across all the underlying assumptions about scabies prevalence in household and age groups, the simple random sampling strategy produces the narrowest 95% confidence interval for all sampling percentages. Based on our simulations of scabies in synthetic populations and the use of different sampling strategies, simple random sampling is more efficient than household or school sampling, as it requires smaller sample sizes and, for some combinations of true prevalence, population size and desired precision, is the only method that requires a sample size smaller than 90% of the population. Compared to random sampling, household sampling requires larger sample sizes to achieve a desired precision. School-based sampling may result in biased estimates of prevalence due to high prevalence of scabies in school-aged children.

When the aim of undertaking a prevalence survey is only to determine whether prevalence is above or below a threshold, then depending on the *a priori* prevalence assumption, high levels of precision may not be required and a smaller sample size may be sufficient. In such cases Table 2 can be used. Where the desired aim of the sample is to determine whether prevalence is above or below a given threshold, say 10%, for the purposes of running a community treatment day: for example, with a medium size population and an *a priori* estimate of true population prevalence between 20–30%, 10% precision would be sufficient to conclude whether the prevalence is higher than 10%. Adopting a simple random sampling strategy, a sample of 5% of the population would be sufficient to reach a decision about whether a community treatment day is required. Note that it is better to overestimate *a priori* prevalence than underestimate, as an underestimate could result in an inadequate sample size.

Simulation approach allows us many scenarios to be investigated [29–35]. In this study, we use a range of population sizes and we test assignment methods consistent with the literature [11, 13–15, 18–20]. Our methodology can be applied to test efficacy of sampling strategies for estimating point prevalence of various infectious diseases. We present a pseudo code as a generic framework to compare sampling strategies in measuring disease prevalence in communities (S5 Fig).

In this work, we do not account for the practicality or cost-effectiveness of undertaking the different types of sampling [3]. In addition, we only consider remote Indigenous communities of Australia with population sizes ranging between 500 and 4000 [36]. Further analysis can be useful to estimate effectiveness of sampling strategies in urban or peri-urban areas with larger population sizes [e.g. 13].

In intervention studies, estimation of prevalence pre- and post-intervention may be necessary [10, 19, 40, 41]. In such cases, a period estimation of disease prevalence, therefore a dynamic transmission model, is needed to accurately represent the impact of intervention on the prevalence [29]. The current version of our approach is not applicable for prevalence estimation in these cases since we only create a snapshot of disease prevalence and test the strategies for estimating point prevalence. As future work, the approach proposed here could be extended to consider sampling populations in pre- and post-intervention periods. In addition, our results show that scabies distribution in communities can provide us some clues about the underlying transmission mechanisms (Fig 4, 5, 6). Our approach can also be extended by comparing these distributions of scabies prevalence in sub-populations to existing survey data of scabies prevalence to infer transmission mechanisms in various populations.

Even though feasibility and cost-effectiveness of sampling strategies are crucial [3], the design of such strategies should take into account the inherent biases that may exist [28, 30, 42]. Due to its feasibility, the WHO recommends school-based sampling to estimate scabies prevalence [28]. However, our results show that the scabies prevalence estimated by using school-based sampling may not be generalisable across the whole community. Our findings highlight the importance of simulation approaches in evaluating and comparing sampling strategies in different population and disease settings.

## Data Availability

Code is available online at https://github.com/nefeltellioglu/sampling_strategy

## Supporting information

**S1 Fig. Observed scabies prevalence in samples for given sampling percentages, across each of the different scabies assignment methods**. The results (median and 2.5% to 97.5% quantiles) are plotted for an exemplar input prevalence percentage between 20–30% across all population sizes where (a) random, (b) high household-specific, (c) mild household-specific, (d) age-specific, (e) age-and-household-specific scabies assignment method is used. Red dashed lines represent 20% and 30% prevalence. Error bars represent the 2.5% to 97.5% quantiles.

**S2 Fig. Observed scabies prevalence in samples selected using different scabies assignment methods and different input prevalence percentages**. The results (median and 2.5% to 97.5% quantiles) are plotted for four exemplar input prevalence percentages of (a) 5%, (b) 10%, (c) 20%, (d) 40% across all population sizes with a sampling percentage of 20%. Red dashed lines represent the input prevalences.

**S3 Fig. Observed scabies prevalence in samples selected using different sampling methods and sampling percentages across populations with (a) small ([500, 1500]), (b) medium ((1500, 2500]), and (c) large sizes ((2500**,**4000])**. The results (median and 2.5% to 97.5% quantiles) are plotted for an exemplar input prevalence percentage between 20-30% with a sampling percentage of 20%. Red dashed lines represent 20% and 30% prevalence.

**S4 Fig. Observed scabies prevalence in samples selected using different sampling methods, sampling percentages, and input prevalence**. The results (median and 2.5% to 97.5% quantiles) are plotted for four exemplar input prevalence percentages of (a) 5%, (b) 10%, (c) 20%, (d) 40% across all population sizes with a sampling percentage of 20%. Red dashed lines represent the input prevalences. In the school-based sampling strategy the highest sampling percentages could not be achieved due to insufficient population size in the school aged group.

**S5 Fig. A generic pseudo code for measuring the efficacy of sampling methods in estimating point prevalence of a given disease**.

**S1 Table. Required sample sizes estimated from simulation results for the simple random, household, and school sampling methods to achieve a given precision, where input scabies prevalence is distributed according to the random method**. Small, medium, and large population sizes represent ranges of [500, 1500], (1500, 2500], (2500, 4000]. For the scenarios with X’s in school sampling, sampling all school-aged children was insufficient to have 95% confidence that prevalence within the selected precision could be obtained.

**S2 Table. Required sample sizes estimated from simulation results for the simple random, household, and school sampling methods to achieve a given precision, where input scabies prevalence is distributed according to the high household-specific method**. Small, medium, and large population sizes represent ranges of [500, 1500], (1500, 2500], (2500, 4000]. For the scenarios with X’s in school sampling, sampling all school-aged children was insufficient to have 95% confidence that prevalence within the selected precision could be obtained.

**S3 Table. Required sample sizes estimated from simulation results for the simple random, household, and school sampling methods to achieve a given precision, where input scabies prevalence is distributed according to the mild household-specific method**. Small, medium, and large population sizes represent ranges of [500, 1500], (1500, 2500], (2500, 4000]. For the scenarios with X’s in school sampling, sampling all school-aged children was insufficient to have 95% confidence that prevalence within the selected precision could be obtained.

**S4 Table. Required sample sizes estimated from simulation results for the simple random, household, and school sampling methods to achieve a given precision, where input scabies prevalence is distributed according to the age-specific method**. Small, medium, and large population sizes represent ranges of [500, 1500], (1500, 2500], (2500, 4000]. For the scenarios with X’s in school sampling, sampling all school-aged children was insufficient to have 95% confidence that prevalence within the selected precision could be obtained.

**S5 Table. Required sample sizes estimated from simulation results for the simple random, household, and school sampling methods to achieve a given precision, where input scabies prevalence is distributed according to the age-and-household-specific method**. Small, medium, and large population sizes represent ranges of [500, 1500], (1500, 2500], (2500, 4000]. For the scenarios with X’s in school sampling, sampling all school-aged children was insufficient to have 95% confidence that prevalence within the selected precision could be obtained.

## Acknowledgments

The household composition data used to supplement Australian Bureau of Statistics data in this paper were collected as part of the Life Course Program. We thank the dedicated Life Course research team who traced participants and collected the data. We especially thank the young adults belonging to the Aboriginal Birth Cohort and their families and community for their co-operation and support and all the individuals who helped in the urban and remote locations. We wish to acknowledge the late Dr. Sue Sayers, founder of the ABC study.

## Footnotes

1 Code is available online at https://github.com/nefeltellioglu/sampling_strategy

