## Supporting Information for "The efficacy of sampling strategies for estimating scabies prevalence"

### Supporting information to manuscript ‘The efficacy of sampling strategies for estimating scabies prevalence’

**S1 Fig. Observed scabies prevalence in samples for given sampling percentages, across each of the different scabies assignment methods.** The results (median and 2.5% to 97.5% quantiles) are plotted for an exemplar input prevalence percentage between 20–30% across all population sizes where (a) random, (b) high household-specific, (c) mild household-specific, (d) age-specific, (e) age-and-household-specific scabies assignment method is used. Red dashed lines represent 20% and 30% prevalence.

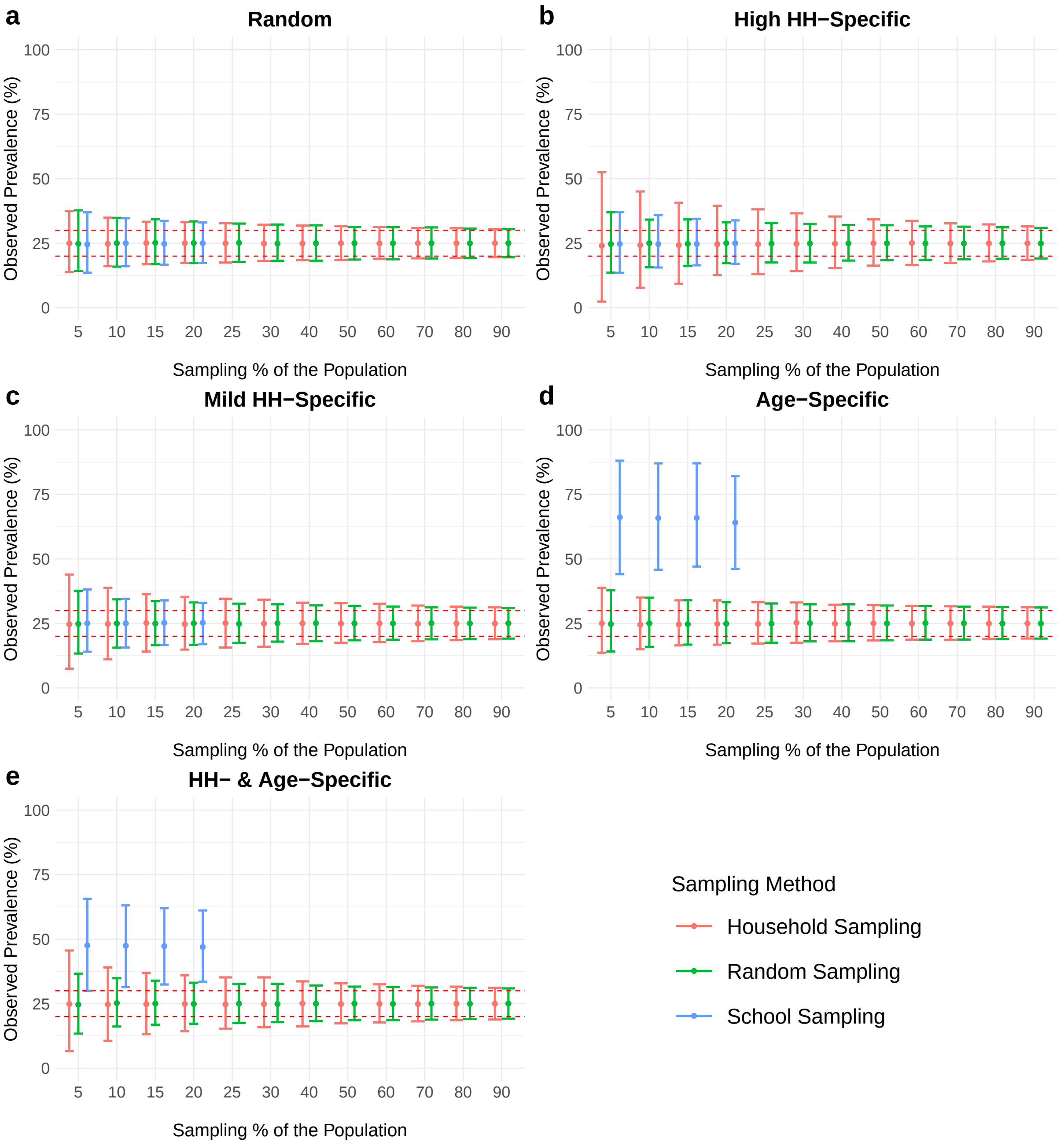

**S2 Fig. Observed scabies prevalence in samples selected using different scabies assignment methods and different input prevalence percentages.** The results (median and 2.5% to 97.5% quantiles) are plotted for four exemplar input prevalence percentages of (a) 5%, (b) 10%, (c) 20%, (d) 40% across all population sizes with a sampling percentage of 20%. Red dashed lines represent the input prevalences.

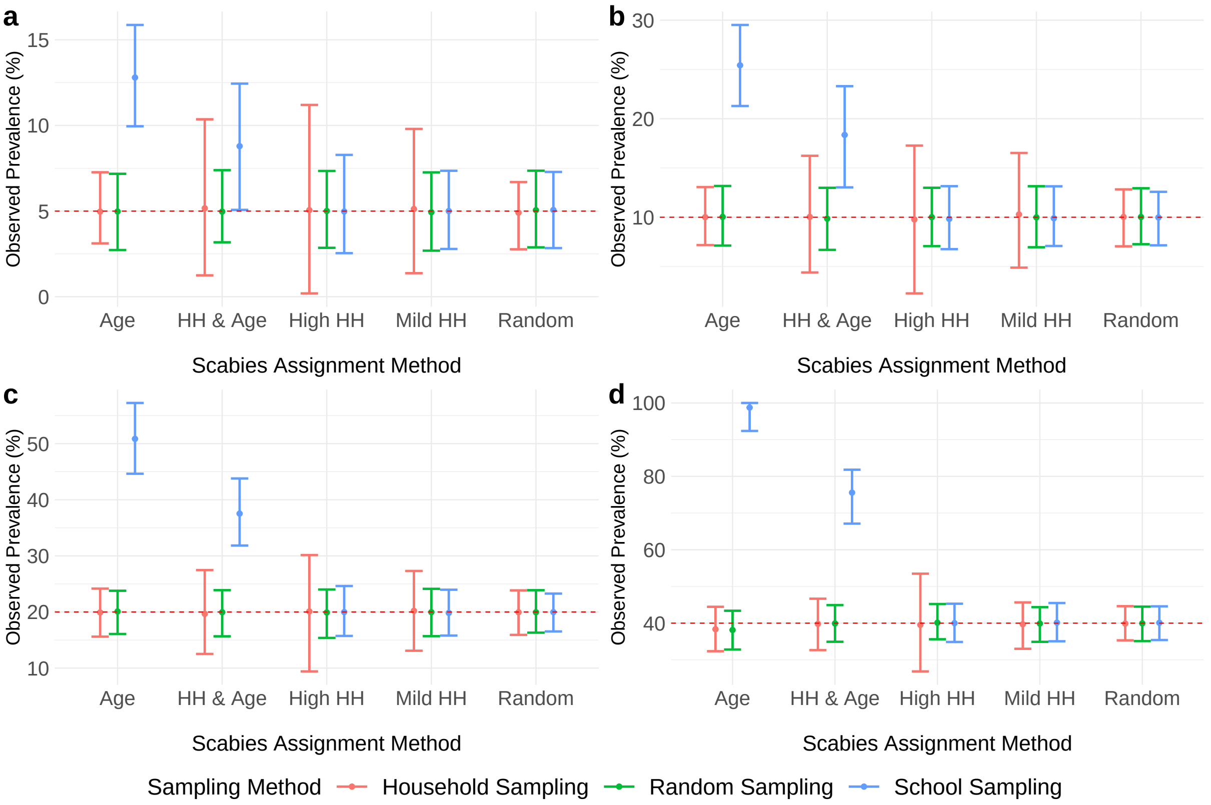

**S3 Fig. Observed scabies prevalence in samples selected using different sampling methods and sampling percentages across populations with (a) small ([500, 1500]), (b) medium ((1500, 2500]), and (c) large sizes ((2500,4000]).** The results (median and 2.5% to 97.5% quantiles) are plotted for an exemplar input prevalence percentage between 20-30% with a sampling percentage of 20%. Red dashed lines represent 20% and 30% prevalence. In the school-based sampling strategy the highest sampling percentages could not be achieved due to insufficient population size in the school aged group.

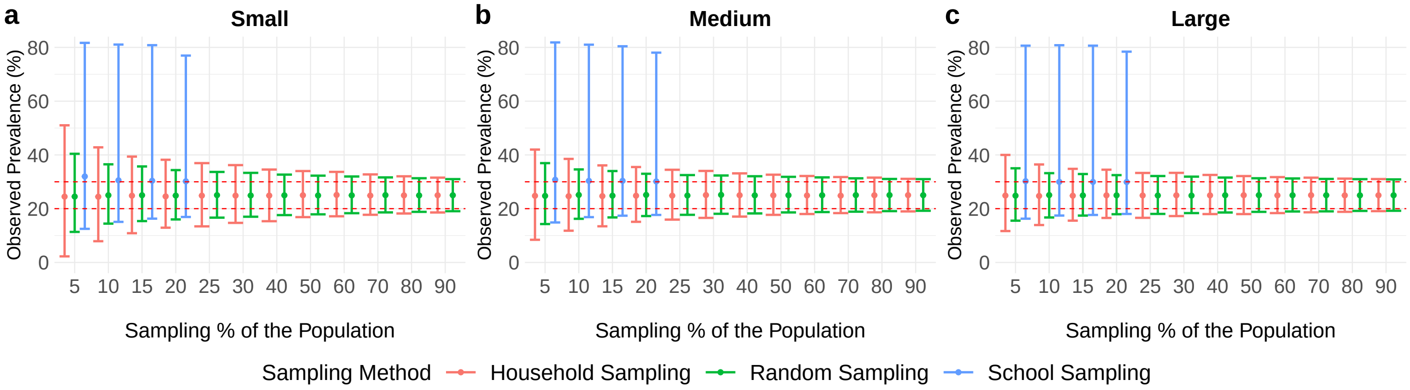

**S4 Fig. Observed scabies prevalence in samples selected using different sampling methods, sampling percentages, and input prevalence.** The results (median and 2.5% to 97.5% quantiles) are plotted for four exemplar input prevalence percentages of (a) 5%, (b) 10%, (c) 20%, (d) 40% across all population sizes with a sampling percentage of 20%. Red dashed lines represent the input prevalences. In the school-based sampling strategy the highest sampling percentages could not be achieved due to insufficient population size in the school aged group.

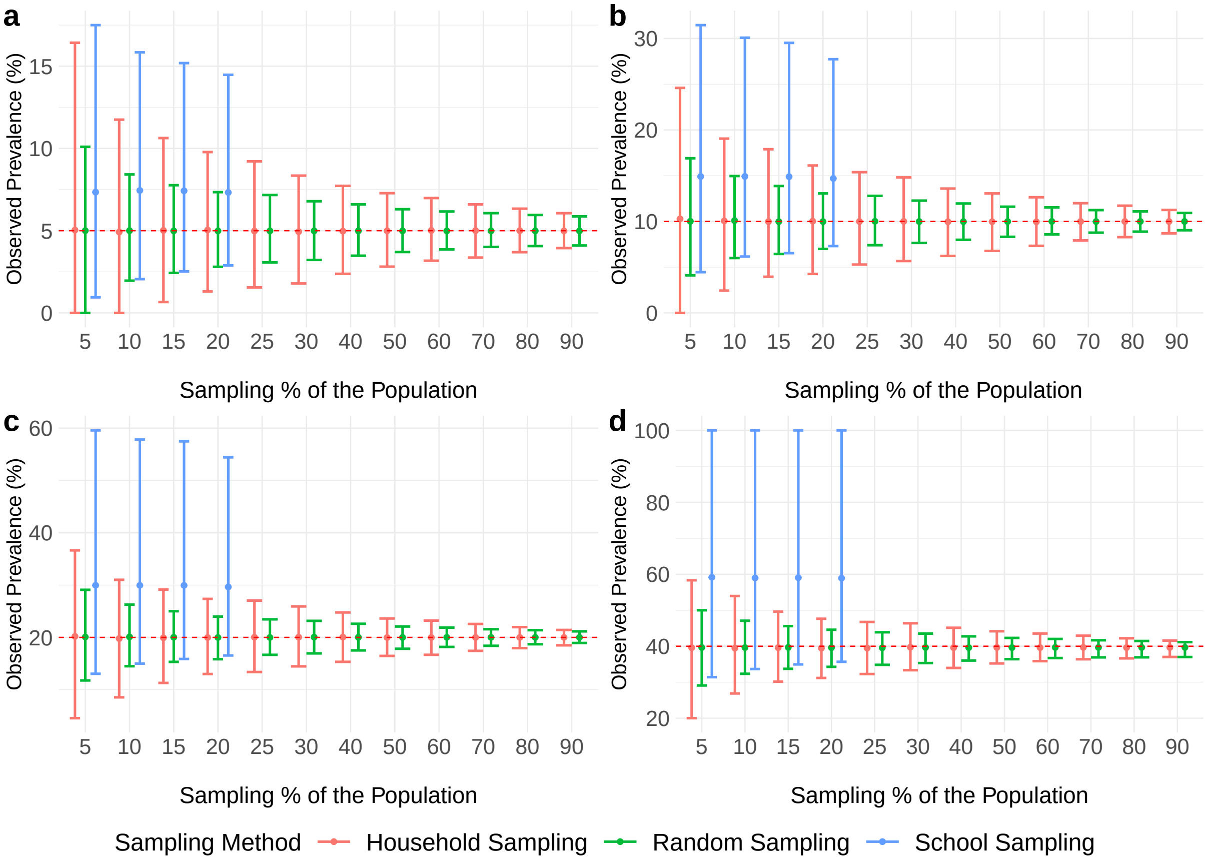

**S5 Fig. A generic pseudo code for measuring the efficacy of sampling methods in estimating point prevalence of a given disease.**

---

Assign population characteristics such as population size ($N$), household distribution, and age distribution in household groups;<br>
Assign *disease assignment methods* based on how disease prevalence differs in sub-populations such as age, gender, and household groups;<br>
Assign a range for *the prevalence percentage* of disease based on *a priori* knowledge of the disease in the population;<br>
Assign a set of *sampling methods* and *sampling percentages*;<br>
Set the number of populations to be generated, $M$;<br>
**for** $i \in \{1, 2, \cdots, M\}$ **do**<br>
    Generate a population dataset, with size of $N$ by assigning household and age groups;<br>
    **for** *each disease assignment method* **do**<br>
        **for** *each specified prevalence percentage* **do**<br>
            Assign scabies to selected individuals based on disease assignment method and specified prevalence percentage;<br>
            **for** *each sampling method* **do**<br>
                **for** *each sampling percentage* **do**<br>
                    Sample the population using the sampling method and sampling percentage;<br>
                **end**<br>
            **end**<br>
        **end**<br>
    **end**<br>
**end**<br>
Compare *the prevalence percentage* in the overall and sampled population

---

**S1 Table. Required sample sizes estimated from simulation results for the simple random, household, and school sampling methods to achieve a given precision, where input scabies prevalence is distributed according to the random method.** Small, medium, and large population sizes represent ranges of [500, 1500], (1500, 2500], (2500, 4000]. For the scenarios with X's in school sampling, sampling all school-aged children was insufficient to have 95% confidence that prevalence within the selected precision could be obtained.

| | | Precision | | | | | | | | |
|---|---|---|---|---|---|---|---|---|---|---|
| | | Simple Random Sampling | | | Household Sampling | | | School Sampling | | |
| Population Size | *a priori* prevalence (%) | 2% | 5% | 10% | 2% | 5% | 10% | 2% | 5% | 10% |
| Small | 5–10 | 50% | 15% | 5% | 50% | 15% | 5% | X | 15% | 5% |
| Small | >10–20 | 60% | 25% | 10% | 70% | 25% | 10% | X | 20% | 10% |
| Small | >20–30 | 80% | 30% | 10% | 80% | 30% | 10% | X | X | 10% |
| Small | >30–40 | 80% | 30% | 15% | 80% | 40% | 10% | X | X | 15% |
| Medium | 5–10 | 30% | 10% | 3% | 25% | 10% | 3% | X | 10% | 3% |
| Medium | >10–20 | 50% | 10% | 5% | 50% | 15% | 5% | X | 15% | 3% |
| Medium | >20–30 | 50% | 20% | 5% | 60% | 15% | 5% | X | 20% | 5% |
| Medium | >30–40 | 60% | 20% | 5% | 60% | 20% | 10% | X | 20% | 5% |
| Large | 5–10 | 20% | 5% | 3% | 25% | 3% | 1% | 20% | 5% | 3% |
| Large | >10–20 | 40% | 10% | 3% | 40% | 10% | 3% | X | 10% | 3% |
| Large | >20–30 | 40% | 10% | 3% | 50% | 10% | 3% | X | 15% | 3% |
| Large | >30–40 | 50% | 15% | 3% | 50% | 10% | 3% | X | 15% | 3% |

**S2 Table. Required sample sizes estimated from simulation results for the simple random, household, and school sampling methods to achieve a given precision, where input scabies prevalence is distributed according to the high household-specific method.** Small, medium, and large population sizes represent ranges of [500, 1500], (1500, 2500], (2500, 4000]. For the scenarios with X's in school sampling, sampling all school-aged children was insufficient to have 95% confidence that prevalence within the selected precision could be obtained.

| | | Precision | | | | | | | | |
|---|---|---|---|---|---|---|---|---|---|---|
| | | Simple Random Sampling | | | Household Sampling | | | School Sampling | | |
| Population Size | *a priori* prevalence (%) | 2% | 5% | 10% | 2% | 5% | 10% | 2% | 5% | 10% |
| Small | 5–10 | 50% | 15% | 5% | >90% | 50% | 20% | X | 15% | 5% |
| Small | >10–20 | 70% | 25% | 10% | >90% | 70% | 40% | X | X | 15% |
| Small | >20–30 | 70% | 40% | 10% | >90% | 80% | 50% | X | X | 10% |
| Small | >30–40 | 90% | 30% | 10% | >90% | 80% | 50% | X | X | 10% |
| Medium | 5–10 | 40% | 10% | 3% | 80% | 25% | 15% | X | 10% | 3% |
| Medium | >10–20 | 60% | 15% | 3% | 90% | 60% | 20% | X | 15% | 3% |
| Medium | >20–30 | 70% | 20% | 5% | >90% | 60% | 25% | X | 20% | 5% |
| Medium | >30–40 | 80% | 20% | 10% | >90% | 70% | 25% | X | X | 10% |
| Large | 5–10 | 30% | 5% | 1% | 70% | 25% | 10% | X | 5% | 3% |
| Large | >10–20 | 40% | 10% | 3% | 90% | 40% | 15% | X | 10% | 3% |
| Large | >20–30 | 50% | 10% | 3% | 90% | 50% | 15% | X | 15% | 5% |
| Large | >30–40 | 50% | 15% | 3% | 90% | 50% | 20% | X | 15% | 3% |

**S3 Table. Required sample sizes estimated from simulation results for the simple random, household, and school sampling methods to achieve a given precision, where input scabies prevalence is distributed according to the mild household-specific method.** Small, medium, and large population sizes represent ranges of [500, 1500], (1500, 2500], (2500, 4000]. For the scenarios with X's in school sampling, sampling all school-aged children was insufficient to have 95% confidence that prevalence within the selected precision could be obtained.

| | | Precision | | | | | | | | |
|---|---|---|---|---|---|---|---|---|---|---|
| | | Simple Random Sampling | | | Household Sampling | | | School Sampling | | |
| Population Size | *a priori* prevalence (%) | 2% | 5% | 10% | 2% | 5% | 10% | 2% | 5% | 10% |
| Small | 5–10 | 60% | 15% | 5% | 80% | 40% | 10% | X | 15% | 5% |
| Small | >10–20 | 70% | 25% | 10% | >90% | 50% | 20% | X | 20% | 10% |
| Small | >20–30 | 80% | 30% | 10% | >90% | 60% | 20% | X | 20% | 10% |
| Small | >30–40 | 90% | 40% | 15% | 90% | 50% | 20% | X | X | 10% |
| Medium | 5–10 | 30% | 10% | 3% | 70% | 25% | 10% | X | 10% | 3% |
| Medium | >10–20 | 60% | 15% | 3% | 80% | 30% | 15% | X | 15% | 5% |
| Medium | >20–30 | 70% | 15% | 5% | 80% | 40% | 15% | X | 15% | 5% |
| Medium | >30–40 | 60% | 20% | 5% | 80% | 40% | 15% | X | X | 5% |
| Large | 5–10 | 20% | 5% | 1% | 60% | 20% | 5% | X | 5% | 3% |
| Large | >10–20 | 40% | 10% | 3% | 70% | 25% | 10% | X | 10% | 3% |
| Large | >20–30 | 60% | 15% | 3% | 80% | 25% | 10% | X | 10% | 3% |
| Large | >30–40 | 50% | 15% | 3% | 70% | 25% | 10% | X | 15% | 3% |

**S4 Table. Required sample sizes estimated from simulation results for the simple random, household, and school sampling methods to achieve a given precision, where input scabies prevalence is distributed according to the age-specific method.** Small, medium, and large population sizes represent ranges of [500, 1500], (1500, 2500], (2500, 4000]. For the scenarios with X's in school sampling, sampling all school-aged children was insufficient to have 95% confidence that prevalence within the selected precision could be obtained.

| | | Precision | | | | | | | | |
|---|---|---|---|---|---|---|---|---|---|---|
| | | Simple Random Sampling | | | Household Sampling | | | School Sampling | | |
| Population Size | *a priori* prevalence (%) | 2% | 5% | 10% | 2% | 5% | 10% | 2% | 5% | 10% |
| Small | 5–10 | 50% | 20% | 3% | 50% | 10% | 5% | X | X | X |
| Small | >10–20 | 70% | 20% | 10% | 80% | 25% | 10% | X | X | X |
| Small | >20–30 | 80% | 30% | 10% | 90% | 40% | 15% | X | X | X |
| Small | >30–40 | >90% | 70% | 10% | >90% | 70% | 15% | X | X | X |
| Medium | 5–10 | 40% | 5% | 3% | 30% | 10% | 3% | X | X | X |
| Medium | >10–20 | 50% | 15% | 5% | 50% | 15% | 5% | X | X | X |
| Medium | >20–30 | 70% | 15% | 5% | 80% | 20% | 10% | X | X | X |
| Medium | >30–40 | >90% | 25% | 5% | >90% | 30% | 10% | X | X | X |
| Large | 5–10 | 20% | 5% | 3% | 20% | 5% | 3% | X | X | X |
| Large | >10–20 | 50% | 10% | 3% | 40% | 10% | 3% | X | X | X |
| Large | >20–30 | 50% | 15% | 3% | 70% | 15% | 3% | X | X | X |
| Large | >30–40 | >90% | 20% | 5% | >90% | 30% | 10% | X | X | X |

**S5 Table. Required sample sizes estimated from simulation results for the simple random, household, and school sampling methods to achieve a given precision, where input scabies prevalence is distributed according to the age-and-household-specific method.** Small, medium, and large population sizes represent ranges of [500, 1500], (1500, 2500], (2500, 4000]. For the scenarios with X's in school sampling, sampling all school-aged children was insufficient to have 95% confidence that prevalence within the selected precision could be obtained.

| | | Precision | | | | | | | | |
|---|---|---|---|---|---|---|---|---|---|---|
| | | Simple Random Sampling | | | Household Sampling | | | School Sampling | | |
| Population Size | *a priori* prevalence (%) | 2% | 5% | 10% | 2% | 5% | 10% | 2% | 5% | 10% |
| Small | 5–10 | 60% | 15% | 5% | 90% | 50% | 20% | X | X | X |
| Small | >10–20 | 70% | 25% | 10% | >90% | 50% | 20% | X | X | X |
| Small | >20–30 | 90% | 25% | 10% | >90% | 60% | 25% | X | X | X |
| Small | >30–40 | 90% | 30% | 15% | >90% | 60% | 20% | X | X | X |
| Medium | 5–10 | 30% | 10% | 3% | 70% | 25% | 10% | X | X | X |
| Medium | >10–20 | 50% | 10% | 3% | 90% | 40% | 15% | X | X | X |
| Medium | >20–30 | 60% | 15% | 5% | 90% | 40% | 15% | X | X | X |
| Medium | >30–40 | 60% | 15% | 5% | 80% | 40% | 15% | X | X | X |
| Large | 5–10 | 30% | 5% | 3% | 60% | 15% | 5% | X | X | X |
| Large | >10–20 | 40% | 10% | 3% | 70% | 25% | 10% | X | X | X |
| Large | >20–30 | 50% | 15% | 3% | 80% | 25% | 10% | X | X | X |
| Large | >30–40 | 50% | 15% | 3% | 80% | 25% | 10% | X | X | X |
